# Exploring the Barriers and Enablers for the Equitable and Accessible Informed Healthcare Consent Process for People with Intellectual Disability: A Systematic Literature Review

**DOI:** 10.1101/2023.03.06.23286791

**Authors:** Manjekah Dunn, Iva Strnadová, Jackie Leach Scully, Jennifer Hansen, Elizabeth Emma Palmer

## Abstract

**Objective:** To identify the factors that act as barriers to, or enablers of, proper informed consent for healthcare interventions for people with intellectual disability.

**Design:** Systematic literature review.

No funding sources or conflicts of interest are reported.

**Data sources:** Databases: Embase, MEDLINE, PsychINFO, PubMed, SCOPUS, Web of Science, and CINAHL (last searched January 2022). Additional articles were obtained from an ancestral search of included articles and hand-searching of three journals.

**Eligibility criteria:** Included studies must examine the informed consent process for a healthcare intervention, be published from 1990 onwards, available in English, and be original research published in a peer-reviewed journal, and participants must be adults and relevant stakeholders (including people with intellectual disability, health professionals, carers or support people, or relevant professionals).

**Synthesis of results:** Inductive thematic analysis using a six-phase method was used to identify factors affecting informed consent. The QualSyst tool was used to assess quality and biases of included studies.

**Results:** Twenty-three studies were included, published from 1999 to 2020, with a mix of qualitative (n=12), quantitative (n=6) and mixed-methods (n=4) studies. Study sizes ranged from 13 to 604 (median 23), and participants included people with intellectual disability, health professionals, carers and support people, and other professionals working with people with intellectual disability. Six themes were identified: health professionals’ attitudes towards and lack of education about informed consent, provision of health information, involvement of carers and other support people, systemic constraints, specific care needs due to patient-related factors, and effective communication between health professionals and patients. Limitations included the heterogeneity of studies, the focus on people with mild intellectual disability only, lack of reflexivity, and limited use of inclusive co-design research methods (n=5).

**Conclusions:** Health professionals’ attitudes and lack of training in informed consent for people with intellectual disability is a major barrier to proper healthcare informed consent for people with intellectual disability. The lack of accessible health information provided for people with intellectual disability also prevents proper informed consent and decision-making. Other factors are the involvement of carers and support people, inherent systemic constraints, failure to meet specific care needs of people with intellectual disability, and ineffective communication by health professionals. Further research, particularly using inclusive co-design methods, is needed to understand these factors. Practical solutions to address these barriers, such as creating accessible information resources and training health professionals, are needed to support improved proper healthcare informed consent for people with intellectual disability.

**Systematic review registration:** PROSPERO number CRD42021290548

## INTRODUCTION

Intellectual disability is a neurodevelopmental condition that affects approximately 1% of the world’s population.[1] The medical definition explains intellectual disability as a group of neurodevelopmental conditions that begin in childhood, characterized by below average cognitive functioning and adaptive behavior, including limitations in conceptual, social, and practical skills.[2] Self-advocates, who are people with intellectual disability who exercise their rights by representing themselves and other people with intellectual disability with whatever supports they need,[3] prefer a different definition. Self-advocates such as Robert Strike OAM (The Medal of the Order of Australia), define intellectual disability in a strengths-based way, highlighting that “Intellectual disability is not an inability to think!” and “We can learn if the way of teaching matches how the person learns”,[4] reinforcing the importance of providing information tailored to the needs of a person with intellectual disability. A diagnosis of intellectual disability, also referred to as ‘learning disabilities’ in the United Kingdom (UK), is associated with significant disparities in health outcomes, including reduced life expectancy compared to the mainstream population.[5-8]

Systemic issues with healthcare delivery systems have resulted in many access barriers for people with intellectual disability, including communication barriers, logistical or programmatic barriers, harmful stereotypes, insufficient accessible health literacy resources and inadequate social support.[9] This is despite direct and indirect disability discrimination legislation, which is in place in many countries who are signatories to the United Nations (UN) Convention on the Rights of Persons with Disabilities.[10] For example, in Australia, the failure of health services and health professionals to make reasonable adjustments during the informed consent process (such as adjusting communication methods to a person’s specific needs, allowing extra time, and providing information in alternative formats including Easy Read options), is an explicit example of both direct and indirect discrimination.[11]

Bodily autonomy, as defined by the United Nations, is an individual’s power and agency to make decisions about their own body, including medical decisions.[12] Informed consent for healthcare – i.e., supporting a person to make health decisions and practice their bodily autonomy – is a fundamental human right outlined by, for example, the National Safety and Quality Health Service Standards (Australia),[13] Mental Capacity Act (UK),[14] and the Joint Commission Standards (United States of America [USA]).[15] Proper practices of informed consent relies fundamentally on three components: 1) the provision of information that the person understands, 2) the decision must be free of coercion, and 3) the person must have capacity.[16] Capacity is the ability to give informed consent for a medical intervention, and for an individual to have capacity, they must be able to understand relevant information, retain the facts, apply the information to make a decision, and communicate that decision.[17, 18] The Mental Capacity Act outlines that “a person must be assumed to have capacity unless it is established that he lacks capacity”, and that incapacity can only be established if “all practicable steps” to support capacity have been attempted without success.[14] Despite these legal protections, patients with intellectual disability continue to be excluded from the medical decision-making process and are not provided the reasonable adjustments that would enable them to give proper informed consent for medical procedures or interventions.[19, 20] This occurs despite evidence that many people with intellectual disability have both the capacity and desire to make their own healthcare decisions.[19, 21] Such exclusion impedes self-determination, which refers to the ability of an individual to act as the causal agents in their lives to make independent decisions.[22] Conversely, improving self-determination for people with intellectual disability has positive impacts on their quality of life, social outcomes, and feelings of empowerment and independence, and can reduce experienced health disparity.[7]

An equitable and accessible informed consent process is required to support people with intellectual disability make independent health decisions.[23] However, people with intellectual disability currently experience inequitable and inaccessible informed consent processes.[19, 24] To address this health gap, we must first understand the factors that contribute to inequitable and inaccessible consent. To the best of our knowledge, the only current review of informed consent for people with intellectual disability is a scoping review by Goldsmith et al. (2008).[25] This review focused on assessments of capacity for informed consent, and noted that the impact of negative attitudes of health professionals and the lack of reasonable adjustments tailoring information to the individual’s needs. However, most of the articles included focus on assessment of capacity [26-28]. More recently there has been a move towards research into ensuring that the consent process itself is accessible for all individuals, for example elderly patients[29] or people with aphasia.[30] However, there remains a paucity of literature on how to optimize the medical informed consent process for people with intellectual disability, and there are currently no systematic reviews summarizing the factors influencing the informed healthcare consent process for people with intellectual disability. Therefore, this study aimed to fill this research gap and identify the barriers and enablers to the informed consent process to best support inclusive, equitable, and accessible healthcare for people with intellectual disability.

### AIMS

A systematic literature review was conducted to examine the literature for papers reporting empirical evidence of factors that act as barriers and enablers to the process of equitable and accessible informed healthcare consent process for people with intellectual disability.

## METHODS

A systematic literature review was conducted following the PRISMA-P systematic literature review protocol.[31] The full study protocol is included in **Appendix 1**.

No patients or members of the public were involved in this research or publication of this manuscript.

### Search strategy

Intellectual disability (and other alternative or historical terminology including ‘learning disabilities’, ‘cognitive disability’, and ‘mental retardation’) was used as the primary search term. Informed consent and its synonyms represented a separate search term (including ‘supported decision-making’, ‘autonomy’, and ‘competence’). Lastly, healthcare interventions were defined as any medical treatment or procedure that aimed to treat, prevent, reduce, or change the natural progression of a disease process.[32]

Multiple databases were searched for articles published between January 1990 to December 2021 (Embase, MEDLINE, PsychINFO, PubMed, SCOPUS, Web of Science, and CINAHL). This yielded 4,853 unique papers which were imported into Covidence, a specialized program for conducting systematic reviews.[33]

### Study selection

Citation screening by abstract and titles was completed by two independent researchers (MD and EP) based on minimum inclusion criteria using Covidence. Included articles needed to:

1. Examine the informed consent process for a healthcare intervention from people with intellectual disability.

2. Have collected most of its data (more than 50%) from relevant stakeholders, including adults with intellectual disability, families, or carers of a person with intellectual disability, and professionals who engage with people with intellectual disability.

3. Report empirical data from primary research methodology.

4. Be published in a peer-reviewed journal after January 1990.

5. Be available in English.

Full text screening was completed by two independent researchers (MD and EP). Articles were excluded if consent was only briefly discussed, or if the study focused on informed consent for research rather than clinical care, assessments of capacity, or participant knowledge or comprehension. Any conflicts were resolved through discussion with an independent third researcher (IS). This screening process identified 21 articles that met inclusion criteria.

Additional studies were identified through an ancestral search and through hand-searching three major journals relevant to intellectual disability research (the British Journal of Learning Disabilities, Journal of Intellectual Disabilities, and Journal of Intellectual Disability Research). Titles were screened and yielded 82 studies which were then screened by abstract. This identified 12 articles then underwent full-text review. This process identified three further articles that met inclusion criteria.

The PRISMA flowchart[34] in **Figure 1** summarizes the study selection.

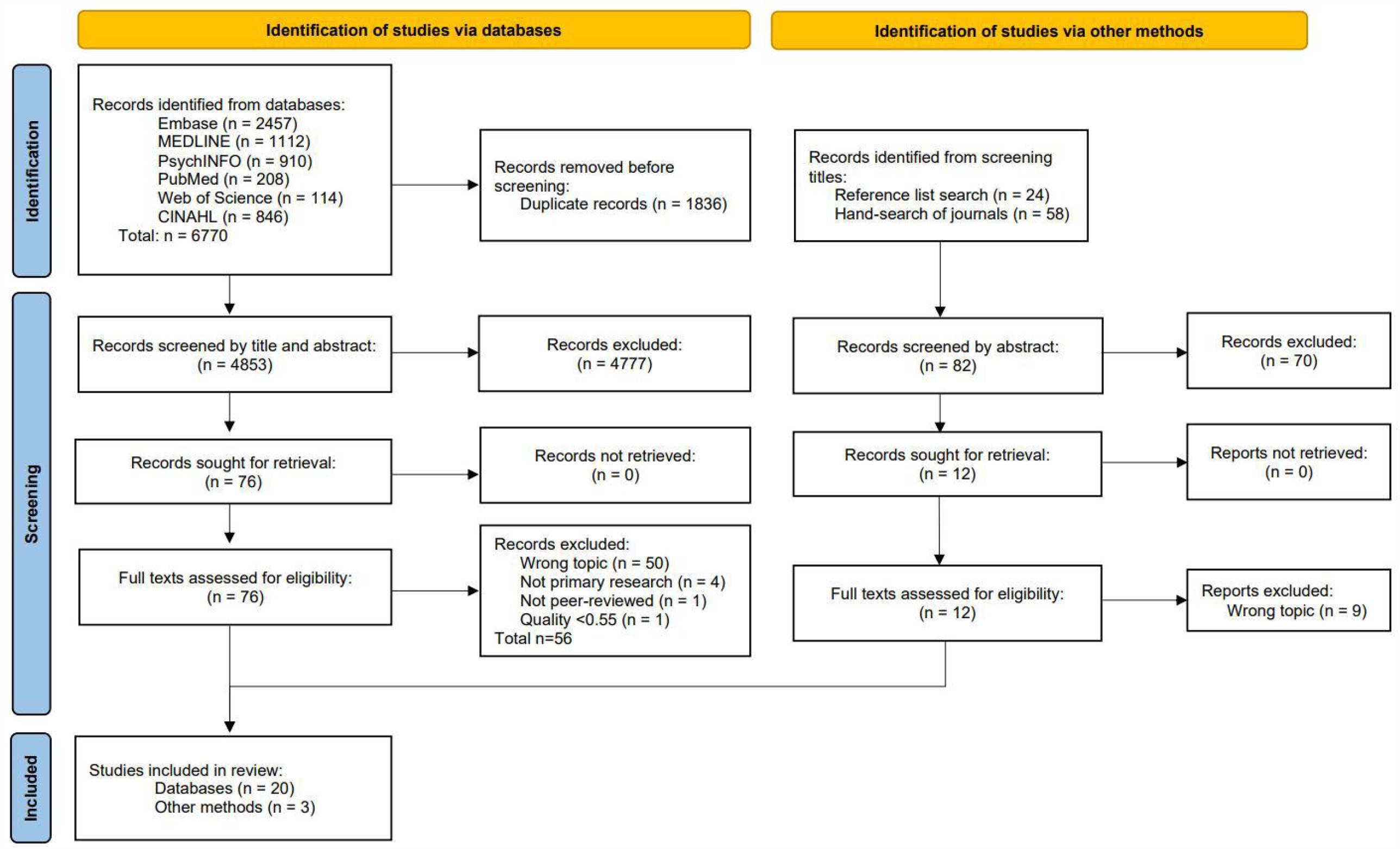

### Quality assessment

Two independent researchers (MD and IS) assessed quality with the QualSyst tool,[35] due to its ability to assess both qualitative and quantitative research papers. After evaluating the distribution of scores, a threshold value of 55% was used, a method suggested by QualSyst[35] to exclude poor-quality studies but ensure sufficient capture of studies overall. Any conflicts between the quality assessment scores were resolved with a third researcher (EP). For mixed-method studies, both qualitative and quantitative quality scores were calculated, and the higher value was used. Common quality limitations identified were a lack of verification procedures to establish credibility, and limited reflexivity of researchers. One article was removed due to a low-quality score of 35%. A total of 23 articles progressed to the data extraction and analysis stage.

### Data collection

The search strategy yielded 23 articles. Two independent researchers (MD and JH) reviewed each study and extracted detailed demographic features, summarized in **Table 1**, including study size, participant demographics, country and year of publication, study design, data analysis, and major outcomes reported. Missing or unclear data were noted, and no assumptions were made. Both researchers (MD and JH) used standardized data collection forms to extract data to address the objectives of this study. These forms specifically sought data on study design, methods, participants, any factors affecting the process of informed consent, and study limitations. Effect size was captured by the volume of text relevant to the outcomes collected, but was not measured in a standardized format due to the heterogeneity of the studies.

**Table 1.**
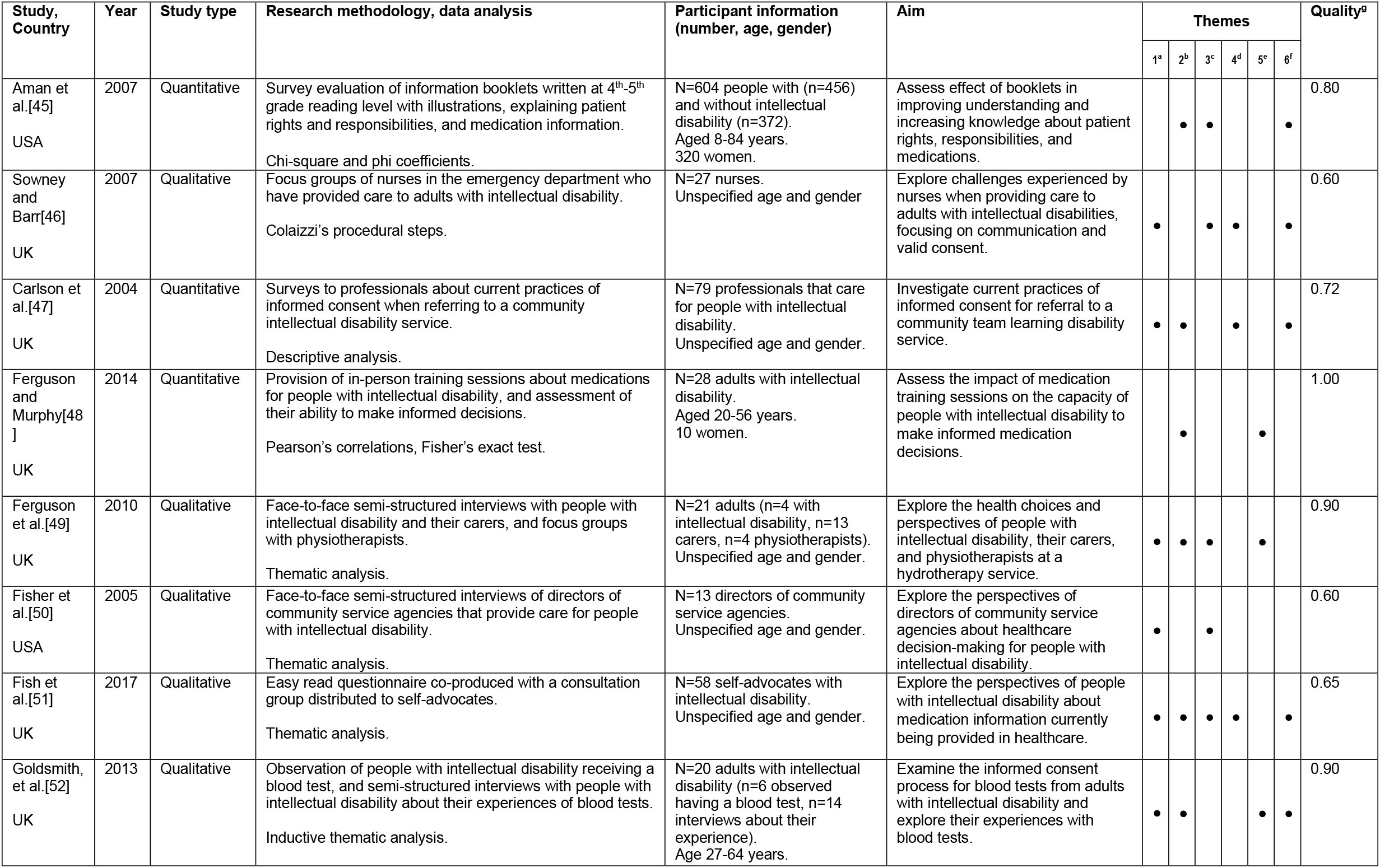
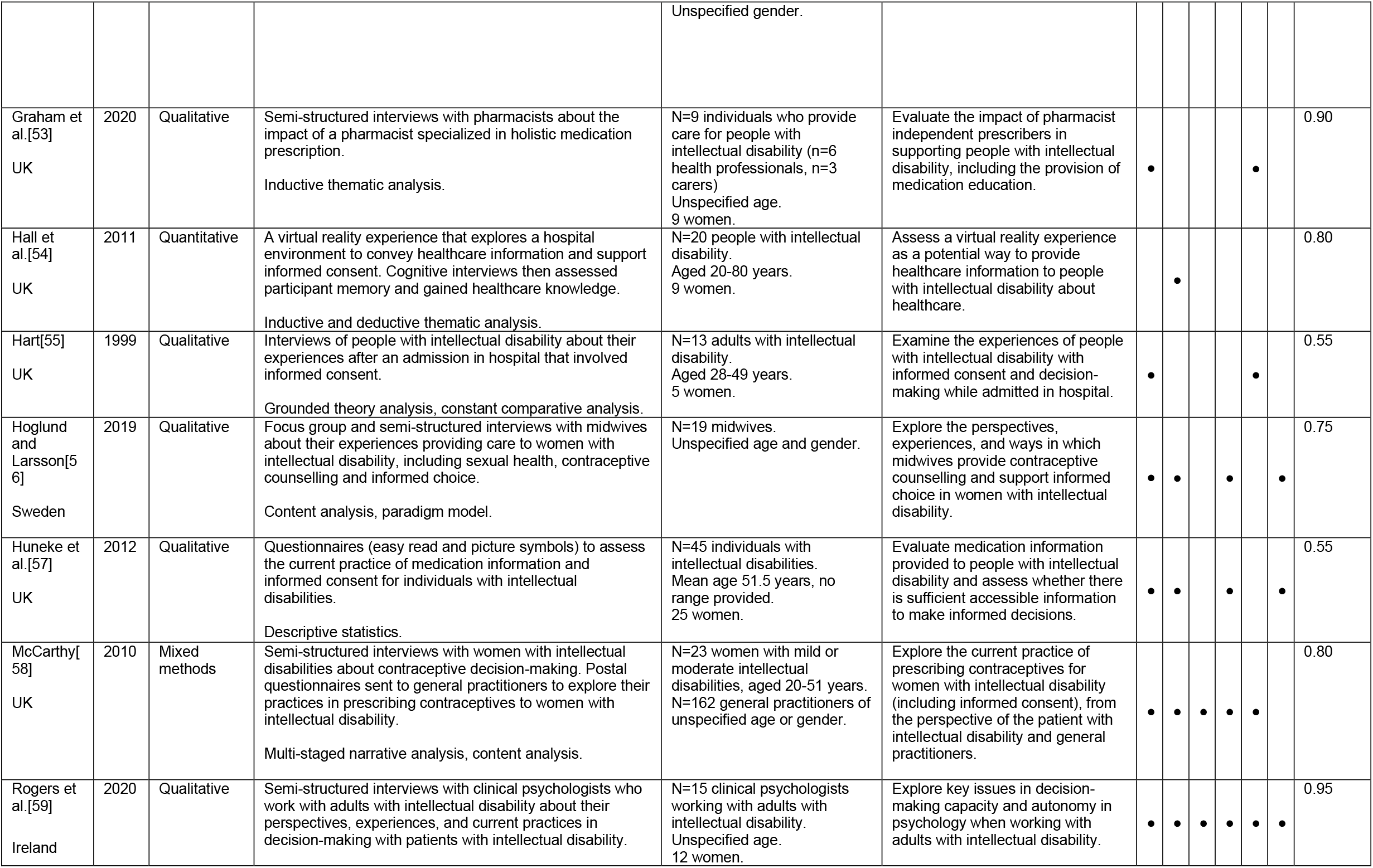
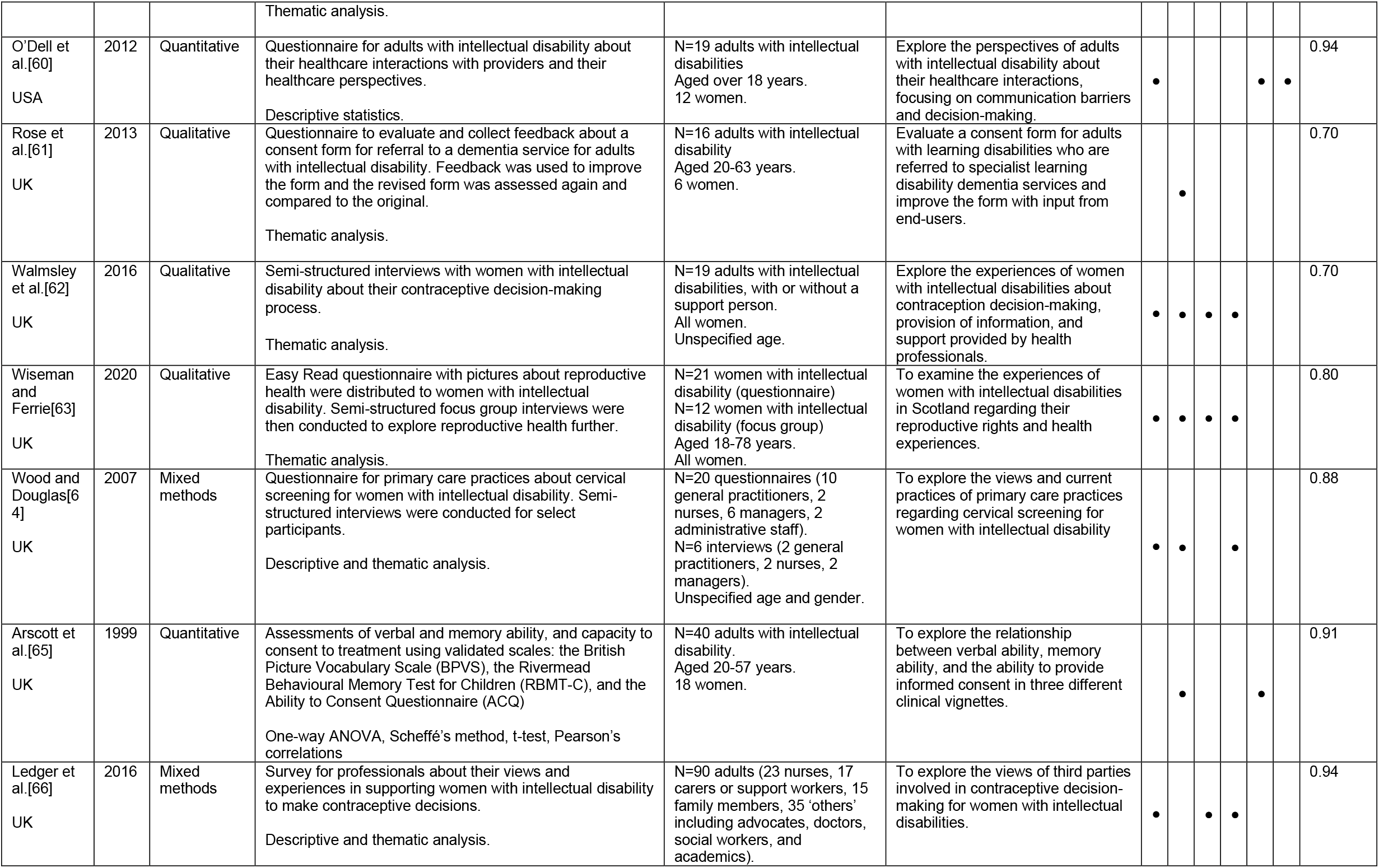
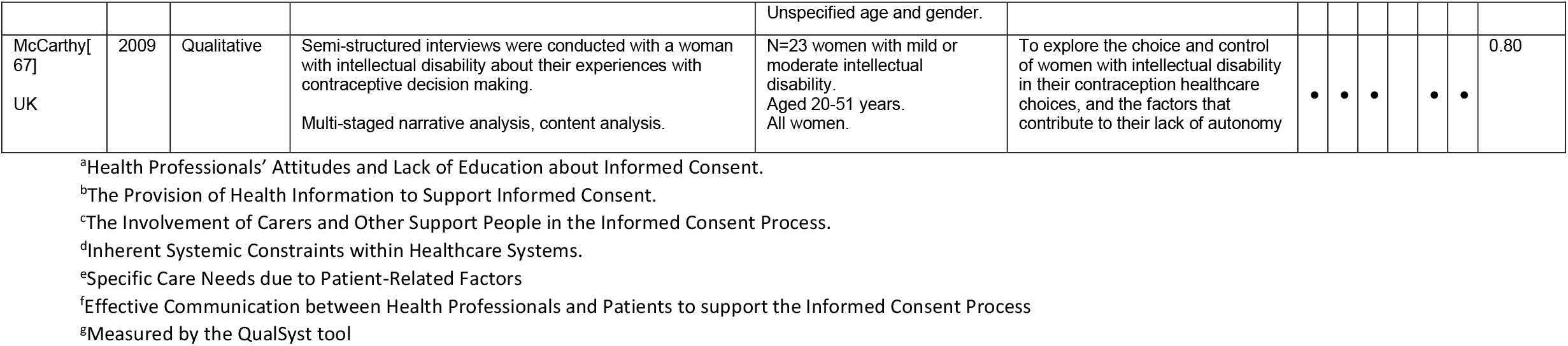
Summary demographic table of included studies, including alignment with the identified six themes.

### Data analysis

The 23 articles had heterogeneous methodology, though the majority contained qualitative data (n=12). Of the quantitative (n=6) and mixed-method (n=4) studies, the majority (80%) used descriptive statistics only. The significant heterogeneity in methodology of the included studies limits the usefulness and applicability of traditional quantitative analysis (i.e., meta-analysis) and inductive thematic analysis is an alternative methodology that can be used in these situations.[36, 37] This method has been previously used in recent systematic reviews examining barriers and enablers to other health processes.[38, 39] No additional processes or methods were required to prepare the data for synthesis. Inductive thematic analysis was completed using the six-phase approach described by Braun and Clarke,[40, 41] consisting of familiarization with data, generation of initial codes, searching for themes, reviewing themes, defining themes, and then writing up the findings. The significance of each theme was assessed using the GRADE framework[42] by considering the quality of studies and their relative contributions.[35] The themes were tabulated in Microsoft Excel to visually display the identified in each study. Study sensitivity was assessed by examining the contributions of included studies to the final themes identified [43].

## RESULTS

Twenty-three articles were identified with a mix of qualitative (n=12), quantitative (n=6) and mixed-methods (n=4) studies. Two papers included the same population of study participants: McCarthy (2009) and McCarthy (2010). However, double counting was not considered an issue in this case as no meta-analysis was completed. Fovargue et al.[44] was excluded due to a low-quality score (35%) and high risk of bias (quality threshold of 55% applied).

Articles were published from 1998 to 2020 and varied in location. Of the 23 selected articles, 18 involved participants from the UK. Three articles had participants from the USA, one from Sweden, and one from Ireland. Study sizes ranged from 13 to 604 (median 23), and participants included people with intellectual disability, health professionals, carers and support people, and other professionals that work with people with intellectual disability (including directors of community service agencies, social workers, and care home staff).

Inductive thematic analysis revealed six major themes which acted as barriers or enablers of the informed consent process for people with intellectual disability, with multiple subthemes identified. The themes were reviewed with two other researchers (EP and IS). **Figure 2** summarizes the identified themes and subthemes.

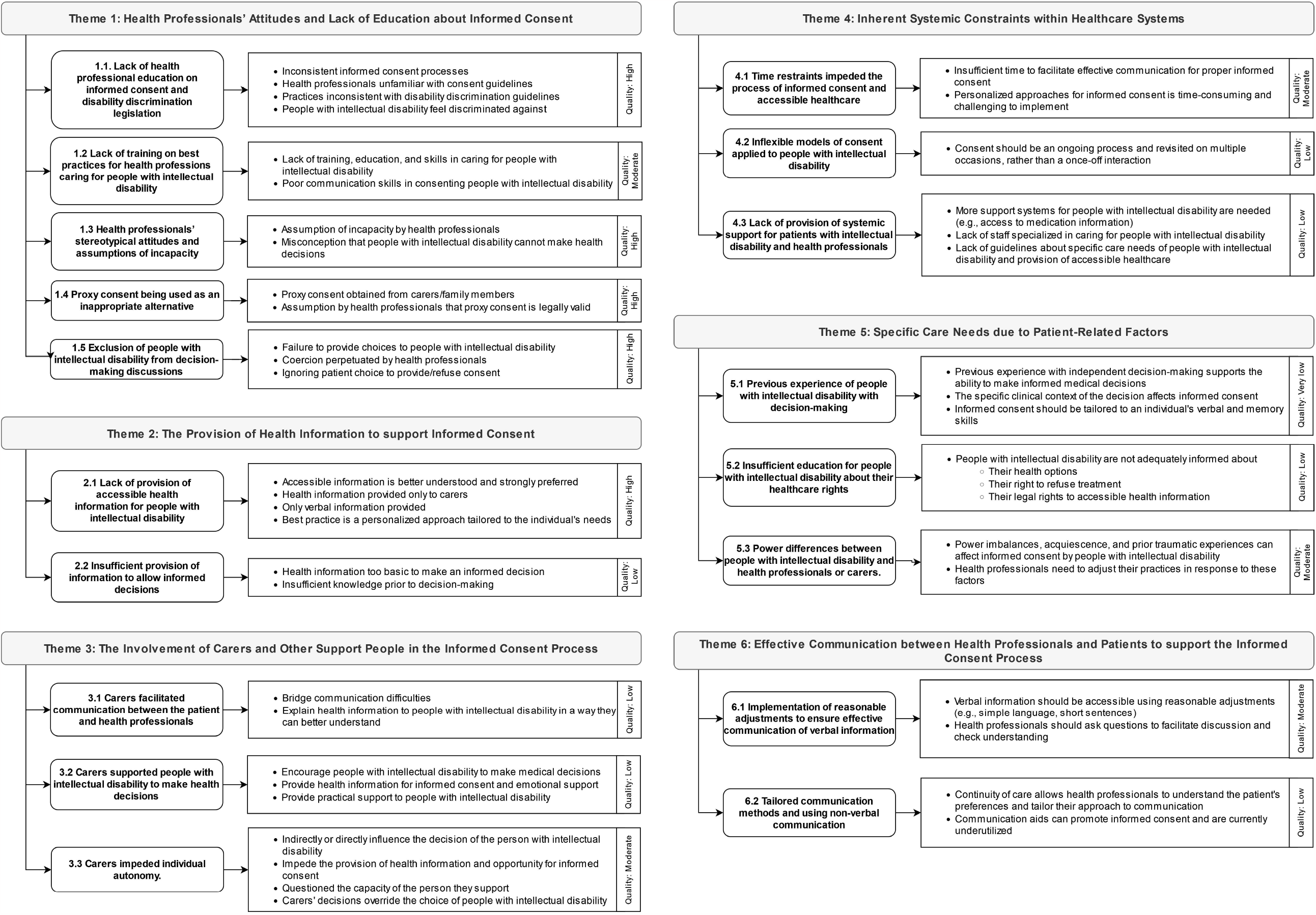

### Theme 1 – Health Professionals’ Attitudes and Lack of Education about Informed Consent

As health professionals are responsible for facilitating proper informed consent for people with intellectual disability, their attitudes and practices were identified as factors affecting the informed consent process in 18 of the 23 articles. Studies reflected the insufficient education and training of health professionals in providing inclusive healthcare to people with intellectual disability, and how ongoing stereotypes and discrimination affect the healthcare they access.

#### 1.1 Lack of health professional education on informed consent and disability discrimination legislation

Inconsistent informed consent practices were described in 12/23 articles[46, 47, 50, 52, 55, 57-60, 62, 66, 67] and reasons for this were multifaceted. While some studies reported that health professionals simply ‘forgot’ to obtain consent or ‘did not realize consent was necessary’,[46, 47] inconsistent consent was also attributed to healthcare providers being unfamiliar with consent guidelines. In a survey by Carlson et al. (2004)[47] only 44% of general practitioners (GPs) stated they were aware of consent guidelines. In this study, reduced awareness of consent guidelines was associated with the misconception that consent from people with intellectual disability was unnecessary. Similar misconceptions were observed in studies of psychologists[59] and nurses[46] where some health professionals felt that informed consent was not necessary for people with intellectual disabilities. Goldsmith et al. (2013) reported inconsistent and poorly informed consent processes during blood tests for people with intellectual disability;[52] they argue that greater training and familiarity with informed consent regulations would improve the consistency of the informed consent process. Furthermore, health professionals surveyed by McCarthy’s study (2010) did not demonstrate awareness of their obligation and responsibility to provide accessible health information to women with intellectual disability.[58] People with intellectual disability feel their health professionals actively discriminated against them because of their diagnosis, reflected in interviews conducted by Wiseman and Ferrie (2020) with comments including “Doctors are useless, and they don’t care about us” and “I can tell, my doctor just thinks I’m stupid – I’m nothing to him”.[63]

#### 1.2 Lack of training on best practices for health professions caring for people with intellectual disability

A lack of health professional training and education in caring for and communicating with people with intellectual disability was also highlighted. A qualitative study of midwives by Hoglund and Larsson [56] reported midwives found it challenging to provide care to people with intellectual disability with limited training. The impact of inadequate skills in communicating with and providing care to people with intellectual disability on the informed consent process was also identified by multiple other health professionals, including psychologists,[59] nurses,[46] pharmacists,[53] and GPs.[58, 64]

#### 1.3 Health professionals’ stereotypical attitudes and assumptions of incapacity

Underlying stereotypes contributed to the belief of health professionals that people with intellectual disability inherently lack capacity and therefore, do not require an attempt at obtaining informed consent. For example, in a survey of health and community professionals referring people with intellectual disability to a disability service, the second most common reason for not obtaining consent was ‘Patient unable to understand’.[47] McGuire et al (2020) interviewed psychiatrists working with people with intellectual disability and many reported on a medical culture that presumes incapacity, where some health professionals appear to have “an unmovable belief” that adults with intellectual disability lack decision-making capacity or see them as “an eternal child”.[59] This is reflected in one participant’s statement: “Some of the challenges are getting across other people’s prejudices… of people with intellectual disability being children who aren’t capable of making any judgement themselves”. Nurses[46] and GPs[58] interviewed in other studies also shared this negative misconception. In a qualitative study of physiotherapists, participants felt there was little to no benefit in providing written hydrotherapy information to people with intellectual disability, and instead perceived their carers as the gatekeepers to influencing healthcare choices.[49]

#### 1.4 Proxy consent being used as an inappropriate alternative

People with intellectual disability are rarely the final decision-maker in most of their medical choices, with many health providers seeking proxy consent from carers, support workers, and family members as alternatives, despite its legal invalidity. Sowney and Barr’s (2007) focus groups with nurses found that many assumed an adult with intellectual disability could not give consent and would seek proxy consent as an acceptable alternative.[46] Similarly, in interviews conducted by McCarthy (2010) of women with intellectual disability, 18/23 stated the decision to commence contraception was made by someone else. Furthermore, the authors’ survey of GPs in the UK found many doctors would gain proxy consent from a carer and did not appear to be aware that proxy consent is invalid in the UK.[58] Overall, this subtheme of proxy consent was identified in 11 of 23 articles in this study.[46, 47, 49-51, 55, 58, 59, 62, 66, 67] Three studies identified that formal documentation of capacity is infrequently completed, which can contribute to an assumption of incapacity and use of proxy consent.[57, 58, 66]

#### 1.5 Exclusion of people with intellectual disability from decision-making discussions

Overall, 5/23 articles described instances where healthcare providers made medical decisions on behalf of their patients with intellectual disability or coerced patients into a choice.[56, 62, 63, 66, 67] Ledger et al. (2016) conducted surveys about contraception decision-making and reported that only 62% of women with intellectual disability were involved in the discussion.[66] The participants with intellectual disability made the final decision in only 38% of cases and the woman’s mother in 16%. In Wiseman and Ferrie’s (2020) study, some women with intellectual disability were not given a contraceptive choice, captured by the comment: “When I went to the doctors to ask about contraception, I was not given the opportunity to explore the different options. I was told what one I should take. I wasn’t encouraged to ask questions or supported to understand all my options”.[63] The authors argue that the lack of effort by health professionals to establish the importance of informed consent prevents these women from meaningfully participating in their own healthcare decisions. Three papers also outlined instances where patient choices were ignored by health professionals, including examples where patients with intellectual disability were denied permission to sign consent forms despite possessing capacity,[50, 55, 59] or a procedure continued despite the patient expressing their wish to withdraw consent.[55]

### Theme 2 – The Provision of Health Information to Support Informed Consent

Proper informed consent relies on the provision of health information to support patient understanding and decision-making, and 17/23 articles identified health information as affecting informed consent for people with intellectual disability. Two major barriers were identified: the lack of accessible health information, and the provision of insufficient information to facilitate informed decision-making.

#### 2.1 Lack of provision of accessible health information for people with intellectual disability

The provision of accessible information was an enabler of the informed consent process in 11 of the 23 included articles.[45, 51, 52, 56-59, 61, 62, 65, 67] In an accessible questionnaire by Fish et al. (2017), self-advocates confirmed their desire for accessible information about their medications, and suggested beneficial features such as large print, pictures, and reducing jargon to support informed medication decision-making.[51] Another study collected feedback from adults with intellectual disability about a medical consent form; comments included reducing the volume of text, using simple language, and including more pictures to improve accessibility.[61] Other identified barriers included health professionals only providing information to carers,[51] not providing Easy Read information due to concerns about ‘offending’ patients,[64] or only giving verbal information.[52] Written accessible information is preferrable, according to semi-structured interviews with women with intellectual disability.[58] Informed consent was supported when health professionals recognized the importance of providing medical information,[49] and when multiple professionals provided this information in an accessible format.[51] Alternative approaches to health information were explored in studies, including virtual reality[54] and in-person education sessions,[48] with varying results. Overall, the need to provide information in different formats tailored to an individual’s communication needs, rather than a ‘one size fits all’ approach, was emphasized by both people with intellectual disability[51] and health professionals.[59]

#### 2.2 Insufficient provision of information to allow informed decisions

Of the included studies, five found that people with intellectual disability were provided with insufficient information to make informed decisions, despite the requirement according to anti-discrimination legislation for health professionals to provide medical information in an accessible format to support the informed consent process.[57, 58, 62, 63, 67] In the study by Fish et al. (2017), 29% of participants with intellectual disability felt they did not receive helpful information from their GP, with the most common reason being that information was too basic.[51] Adults with intellectual disability surveyed by Rose et al. (2013) expressed their desire for additional information to improve a consent form for referral to a dementia service.[61] In McCarthy’s (2009) study, some women with intellectual disability believed the contraceptive pill was only prescribed to control menstruation and they were not made fully aware that it prevented pregnancy, raising concerns that their ‘consent’ was thus not truly informed.[67]

### Theme 3 – The Involvement of Carers and Other Support People in the Informed Consent Process

Support people (including carers, family members, and group home staff) were identified in 11 articles as affecting the informed consent process. There were varying results, with some studies emphasizing the role of carers in supporting and promoting decision-making for a person with intellectual disability,[45, 46, 51, 62, 66] while other studies[50, 59] highlighted that carers also obstruct or prevent the informed consent process, and a few studies reflected both aspects.[49, 58, 63, 67]

#### 3.1 Carers facilitated communication between the patient and health professionals

Carers who are familiar with the communication needs of the person with intellectual disability can bridge communication difficulties to promote informed consent.[46, 58] Ferguson et al. (2011) reported that carers felt that they could recognize subtle communication cues to understand the preferences of the person with intellectual disability.[49] McCarthy’s study (2010) interviewed women with intellectual disability and found that 21/23 women preferred to see doctors with a support person due to their perceptions of communication benefits: “Sometimes I don’t understand it, so they have to explain it to my carer, so they can explain it to me easier”.[58] Most GPs surveyed in this study (93%) also stated that a support person aided communication.

#### 3.2 Carers supported people with intellectual disability to make health decisions

By acting as advocates of people with intellectual disability, carers can encourage decision-making,[49, 63] provide health information,[63, 66] provide emotional support, and assist with practical tasks such as reading health information or remembering important information.[45, 62, 67] Some people with intellectual disability explicitly appreciated their support person’s involvement,[51] such as in McCarthy’s study (2009) where 18/23 participants perceived a support person as advantageous, and felt supported and safer when a support person was involved.[67]

#### 3.3 Carers impeded individual autonomy

Although McCarthy’s study (2009) found 21/23 women with intellectual disability did not feel comfortable seeing a doctor alone to discuss contraception, the study argued that a support person could exclude the woman with intellectual disability from the discussion, and that the woman’s choice could be influenced by the carer’s presence.[67] Wiseman and Ferrie’s (2020) study found that while younger participants with intellectual disability stated family members empowered their decision-making, older women felt family members impaired their access to informed consent: “Your mum and dad don’t want to tell you about things like that because they think because you’ve got a learning disability you don’t understand! They hide it away from you because it’s better for them”.[63] Carers interviewed by Ferguson et al. (2011) questioned the capacity of the person with intellectual disability they supported, and stated they would guide the person with intellectual disability to pick the ‘best choice’, or in some cases would over-ride the choices of the person with intellectual disability.[49] In a study of psychologists,[59] participants described instances where the decision of family or carers was prioritized over the wishes of the person with intellectual disability, and this was also seen in another study by Fisher et al. (2005).[50] This prioritization directly impedes the personal autonomy of the person with intellectual disability: and indeed, people with intellectual disability expressed their frustration when others become involved in decision-making about their lives.[49] McCarthy (2010) also argued that parents could coerce medical choices, for instance where women with intellectual disability feared pregnancy and felt pressured to start contraception because “My dad wouldn’t like it” or in some cases were not given the opportunity of choice: “My mum doesn’t want me having babies, so she got me to use the injection”.[58] Other women interviewed by McCarthy (2009) feared they would lose their group home support or familial support if they became pregnant, and consequently felt that they had no choice, reflected in one participant’s statement “she said they’d get rid of me if I was pregnant”.[67]

### Theme 4 – Inherent Systemic Constraints within Healthcare systems

Systemic constraints in current healthcare models were identified in 9/23 articles and acted as barriers to proper informed consent. Common subthemes included time restraints, inflexible models of care, and insufficient supports to both people with intellectual disability and health professionals.

#### 4.1 Time restraints impeded the process of informed consent and accessible healthcare

Five of the 23 included studies identified that resource limitations of current healthcare models create time constraints which impaired the equitable and accessible consent process for patients with intellectual disability.[46, 47, 58, 59, 64] This was identified as a barrier in qualitative studies of psychologists,[59] GPs,[58] hospital nurses,[46] and in community disability workers.[47] McGuire et al. (2020) argued the inflexible medical models in Ireland create time restraints that impede informed decision-making, with suggested solutions including hospital passports (i.e., a document containing key information regarding an individual’s needs, given to health professionals during hospital presentations,[68] person-centered plans, and multimedia technologies.[59] Unfortunately, this personalized approach to informed consent is difficult to implement practically and only two studies described instances where health professionals supported the informed consent process by demonstrating flexible personalized approaches, for example by providing multimodal information tailored to the individual,[51] using adaptive approaches to consent depending on patient needs,[56] and longer appointment times.[51, 56] A survey of primary care practices reported that most do not modify their cervical screening information to make it accessible to patients with intellectual disability, stating it was more practical to send the same information to all patients rather than individualize it for their needs.[64]

#### 4.2 Inflexible models of consent applied to people with intellectual disability

Another subtheme involved the current models of informed consent processes. Both people with intellectual disability[62] and health professionals[59] recognized that consent is traditionally obtained through one-off interactions prior to an intervention, but for people with intellectual disability consent should ideally be an ongoing process that begins before an appointment and continues between subsequent ones. Other studies that observed informed consent tended to describe one-off interactions where decision-making was not revisited at subsequent appointments.[51, 56, 58]

#### 4.3 Lack of provision of systemic support for patients with intellectual disability and health professionals

A survey of self-advocates highlighted the lack of support systems for people with intellectual disability to improve their health.[51] They suggested a telephone helpline and a centralized source of information so that people with intellectual disability can access information about their medications. Health professionals also felt a need for greater systemic supports when seeing patients with intellectual disability. Hoglund and Larsson (2019) reported midwives wanted health professionals specialized in intellectual disability care to provide support for other staff when caring for this patient group.[56] Another study found benefit in having a pharmacist specifically to support patients with intellectual disability with their medications.[53] These issues are compounded by an ongoing lack of accepted guidelines to guide health professionals about the specific care needs of people with intellectual disabilities, such as in contraceptive counselling[56] or primary care.[64]

### Theme 5 – Specific Care Needs due to Patient-Related Factors

Ten studies (of 23) identified patient-related factors that influenced proper informed consent for people with intellectual disability. These were grouped into three main subthemes: previous experience with and skills in decision-making, insufficient education about healthcare rights, and power differences.

#### 5.1 Previous experience of people with intellectual disability with decision-making

A study by Arscott et al. (1999) concluded that an individual’s verbal and memory skills affected their capacity to consent, and that if health information is made less dependent on verbal or memory ability then informed consent can be promoted.[65] In this study, participants’ ability to consent also changed with different clinical vignettes, supporting the view of ‘functional’ capacity specific to the context of the medical decision. Although previous experiences with decision-making did not influence informed consent in this paper, other studies have suggested that people with intellectual disability who are accustomed to independent decision-making are more able to make informed medical decisions.[59, 60] Individuals with intellectual disability who live independently were more likely to make independent healthcare decisions, compared to those who lived with parents or carers.[58]

#### 5.2 Insufficient education for people with intellectual disability about their healthcare rights

Another subtheme identified in five of the 23 included studies[49, 52, 58, 65, 67] was that people with intellectual disability were not adequately informed that they have health choices or that they can refuse treatment. This raises concerns about whether consent is truly ‘informed’. In some cases, such as in the study by Ferguson et al. (2011) about the decision to attend hydrotherapy,[49] medical decisions were not presented to people with intellectual disability as a choice, and therefore, they do not perceive they have had an opportunity to express their autonomy. Furthermore, women with intellectual disability interviewed in McCarthy’s study (2010) appeared to be uninformed of their legal right to accessible health information.[58]

#### 5.3 Power differences between people with intellectual disability and health professionals or carers

Acquiescence by people with intellectual disability – i.e., their tendency to agree with suggestions made by carers and health professionals, often to avoid disagreements or upsetting others due to common and repeated experiences of trauma – was identified in five studies as an ongoing barrier to the informed consent process.[52, 53, 55, 59, 67] In McCarthy’s (2009) interviews of women with intellectual disability, some participants implicitly rejected the idea that they might make their own healthcare decisions, reflected in a participant’s statement: “They’re the carers, they have responsibility for me.”, while other women appeared to have made decisions to appease their carers: “I have the jab (contraceptive injection) so I can’t be blamed for getting pregnant”.[67] Previous traumatic experiences also influenced some participant’s medical choices: “What’s point of having a baby, when you’ve already had one taken away?” and McCarthy emphasised the importance of providing trauma-sensitive care to people with intellectual disability. To support the proper informed consent process, two studies highlighted that health professionals need to be mindful of power imbalance when providing care for people with intellectual disability to ensure the choices truly reflect autonomy.[53, 59]

### Theme 6 – Effective Communication between Health Professionals and Patients to support the Informed Consent Process

Communication is a key component of informed consent as information must be provided accurately, and the patient’s decision must also be conveyed effectively. Informed consent can be affected by ineffective communication, and this was identified in 8/23 articles and reflected by both people with intellectual disability and health professionals.

#### 6.1 Implementation of reasonable adjustments to ensure effective communication of verbal information

In general, simple language and avoiding complicated words in verbal communication was preferred by people with intellectual disability.[51, 52] Hoglund and Larsson (2019) interviewed midwives and found that they used a variety of communication aids and approaches to facilitate medical decision-making, including repetition, short sentences, models, pictures, and Easy Read brochures.[56] In a questionnaire to self-advocates about medication consent conducted by Fish et al. (2017),[51] participants with intellectual disability emphasized their preference for clear, simple verbal information, for example: “Any info in a verbal equivalent of easy-read, plain English would help”. In McCarthy’s (2009) study, women with intellectual disability interviewed about contraceptive decision-making reported the absence of question-asking and discussion when making medical decisions, recalling that they themselves did not ask questions as the patient, and the health provider also did not ask any questions before a decision was made.[67]

#### 6.2 Tailored communication methods and using non-verbal communication

Midwives providing contraceptive care for women with intellectual disability highlighted that continuity of care allows health professionals to develop rapport and understand the communication preferences of people with intellectual disability to facilitate the informed consent process.[56] This is not always possible in medical contexts; for emergency nurses encountering a patient with intellectual disability for the first time, the lack of background patient information about patients with intellectual disability made it challenging to understand their communication preferences within the fast-paced Emergency Department environment.[46] People with intellectual disability in one study also supported the use of hearing loops, braille, and sign language.[51] The use of non-verbal communication, such as body language, was also highlighted as an underutilized aspect of the current medical model of consent and patient communication by Huneke et al. (2012)[57] and McGuire et al. (2020).[59]

## DISCUSSION

To the best of our knowledge, this is the first systematic review investigating the barriers and enablers of the informed consent process for healthcare procedures for people with intellectual disability. Since the 2000s, there has been a paradigm shift in the way informed consent and capacity is viewed, moving away from the idea that capacity is a fixed ability that only some individuals possess. Instead, capacity is viewed as ‘functional’: a flexible ability that changes over time and in different contexts.[69] This implies that deficits in capacity can be improved by using accessible informed consent resources, or training and educational interventions.[18] In most countries health professionals have the mandatory requirement to maximize the patient’s ability to consent through reasonable adjustments and effective communication.[13] By acknowledging the barriers and enablers identified in this review, physicians can best support the informed consent process for their patients with intellectual disability.

This review has found the provision of healthcare to people with intellectual disability is inequitable and inaccessible due to poor informed consent practices. The studies included reflect a lack of respect for personal autonomy for people with intellectual disability, partly due to continuing assumptions of incapacity and negative stereotypes, despite disability discrimination legislation protecting their rights to healthcare.[10] These beliefs are perpetuated by health professionals and carers and are reflected in the incorrect use of proxy consent as an alternative, despite its legal invalidity. Health providers and carers that assume incapacity of people with intellectual disability deny this population their bodily autonomy and patient right to give informed consent. In some studies, the choices of people with intellectual disability were ignored completely, and many are not given the opportunity to make informed decisions. This is compounded by inadequate education of health professionals about consent guidelines and discrimination legislation, and ineffective communication with patients with intellectual disability. Our findings reflect the broader systemic barriers of resource limitations and inadequate staff training in providing care to people with intellectual disability that contribute to the ongoing shortfalls of the current medical system and poor health outcomes for people with intellectual disability. As health professionals are the primary gatekeepers to proper informed consent, it is essential that medical systems provide adequate education and training to support them in working collaboratively with their patients with intellectual disability to enable proper informed consent processes. Part of this responsibility also includes informing patients with intellectual disability of their legal rights to autonomy and accessible information and supporting them to advocate for themselves and their own patient rights. Adequate education and training for health professionals is urgently needed to address poor informed consent practices and this should be supported by inclusive resources for people with intellectual disability.

This study also found a lack of resources to support equitable and accessible informed consent processes within the current medical model. As informed consent relies upon the provision of sufficient healthcare information to support decision-making, it is unsurprising that the way health professionals provide information is highly relevant. Almost half of the included studies (11/23) described the shortage of accessible, Easy Read medical information to support effective communication and medical decision-making. All patients, including people with intellectual disability, have the legal right to accessible health information to make informed decisions, with protections including the UK’s Accessible Information Standard,[70] and Australia’s Charter of Healthcare Rights.[23] These documents outline that patients have the right to receive information in a format they understand, and communication support should be provided if required to make an informed choice. Although policy regulations exist, there is ongoing evidence that these are not regularly observed for patients with intellectual disability. The current literature explores a variety of ways to address this information barrier, but the heterogeneity of studies makes it challenging to generalize about what specifically makes health information ‘accessible’. However, this study emphasizes the importance of using simple language (ideally in an Easy Read format), multimodal resources, and a patient-centered flexible approach to the provision of health information to support the process of informed consent. With a welcome focus on patient-centered decision-making and autonomy, there is an urgent need for inclusive informed consent practices to ensure people with intellectual disability are equally able to access modern medicine. For example, the growing field of genetic medicine relies heavily on the proper informed consent process with substantial pre- and post-test counselling, and the current inequitable and inaccessible informed consent model effectively excludes people with intellectual disability from receiving inclusive genetic testing.[19]

Limitations of the literature include the scarcity of inclusive research practices in most studies involving people with intellectual disability as participants, making results vulnerable to external biases, such as in the use of inaccessible questionnaires for people with intellectual disability, involvement of carers in the data collection process, absence of reflexivity by researchers, and lack of consideration regarding the effects of over-compliance on research findings. The use of advisory groups or co-research with people with intellectual disability are ways to promote inclusive research but were only used in five studies. Furthermore, studies often had vague selection criteria, low sample sizes, missing data, and used various gatekeepers to identify eligible participants with intellectual disability which increases the risk of bias. The QualSyst tool evaluates this through assessments of reflexivity and use of triangulation to improve result validity, and this was considered in the assessment of study results and data analysis. Of the 15 studies that included people with intellectual disability as participants, only one study specifically focused on people with intellectual disability with higher support needs, and nine studies explicitly excluded those with severe or profound intellectual disability. This reflects an ongoing selection bias in intellectual disability research, which tends to focus on people with mild intellectual disability. There is a continuing need for intellectual disability research that reflects the voices of people with intellectual disability, and research that uses inclusive practices. In particularly, more needs to be known about barriers to informed consent from the perspective of people with intellectual disability, and attention should also be placed on training and educating health professionals to provide proper informed consent to achieve equitable and accessible healthcare for people with intellectual disability. Limitations to the review process were related to the heterogeneity of studies, which made it challenging to apply meta-analysis and directly compare findings. The variation in study design and highly qualitative nature of studies meant that effect sizes could not be easily applied. Qualitative methods of effect size calculations have been described in the literature [71, 72] and effect measures were captured by evaluating the volume of text and amount of evidence in each article dedicated to the six identified themes, but the heterogeneity in study design and outcomes meant it was challenging to apply a standardized method as it would not accurately reflect the true effect size. The qualitative nature of the studies also made traditional sensitivity analysis challenging. Nonetheless, after thematic analysis was completed, the relative contributions of each study were examined, guided by previously described methods for qualitative reviews [43]. Higher-quality studies, and studies that included people with intellectual disability as participants, generally contributed more significantly to the findings of this study.

## CONCLUSION

This review examined the barriers and enablers of the healthcare informed consent process for people with intellectual disability. Poor and inconsistent consent practices of health professionals and the lack of accessible health information emerged as two key themes affecting the informed consent process, highlighting the need for better training of health professionals and accessible health information specifically for people with intellectual disability. Other contributing factors include the role of support people and carers, systemic healthcare problems, patient-related aspects, and challenges in effective communication between health provider and patients. A multisystem approach is needed to address these factors and shift the current consent model towards one that is more inclusive and supportive of people with intellectual disability. Future research must address these barriers to the informed consent process through inclusive research practices, such as interviews with people with intellectual disability and with health professionals, and additional studies are needed about consent in specific contexts such as cancer screening and treatment, obstetrics and gynecology, psychiatry, and genomic medicine. For example, our inclusive GeneEQUAL program has found a lack of studies regarding inclusive informed consent for people with intellectual disability about genetic testing,[21] and evidence of exclusionary and disrespectful practice by genetic professionals,[19] despite a growing number of genetic tests being performed on people with intellectual disability,[73] and the opening of several advanced therapeutic trials such as gene-based therapies for genetic causes of intellectual disability.[74]

## Data Availability

All data produced in the present study are available upon reasonable request to the corresponding author.

